# Ticagrelor responsive platelet genes are associated with platelet function and bleeding

**DOI:** 10.1101/2022.03.18.22270280

**Authors:** Rachel A Myers, Thomas L Ortel, Alexander Waldrop, Sandeep Dave, Geoffrey S Ginsburg, Deepak Voora

## Abstract

**Importance:** Ticagrelor inhibits platelet function, prevents myocardial infarction, and causes bleeding. A comprehensive analysis of the on- and off-target platelet effects of ticagrelor that underlie its clinical effects is lacking.

**Objective:** To test the hypothesis that platelet transcripts that change in response to ticagrelor exposure are associated with platelet function or bleeding.

**Design:** A discovery cohort of healthy volunteers were sequentially exposed to aspirin, aspirin washout, and ticagrelor. Messenger RNA sequencing (mRNAseq) of purified platelets was performed pre/post each exposure. We defined the ticagrelor exposure signature (TES) as the ratio of mean expression of up-vs. down-regulated genes by ticagrelor that were prioritized based on lasso regression, weighted gene co-expression networks, and isoform level analyses. A separate healthy cohort was recruited to validate ticagrelor’s effects on TES genes measured using Nanostring. Platelet function was measured at baseline and in response to ticagrelor exposure in all participants. Self-reported bleeding was systematically queried during periods of ticagrelor exposure.

**Setting:** An early phase, academic, clinical research unit.

**Participants:** Self-reported, healthy volunteers age > 30 and < 75, non-smoking, taking no daily prescribed medications.

**Exposures:** Ticagrelor (90 mg twice daily) and aspirin (81 mg/day and 325 mg/day) each for 4 weeks.

**Main outcomes and measures:** Expression levels of platelet messenger RNA, platelet count, mean platelet volume, and 9 different measures of *ex vivo* platelet function (aggregated into a previously described platelet function score), and self-reported bleeding at baseline and after each exposure.

**Results:** In the discovery cohort (n = 58, mean age 43, 39 female) platelet mRNAseq identified (FDR < 5%) 1820 up- and 1589 down-regulated genes associated with ticagrelor exposure. We prioritized 84 of these transcripts to calculate a TES score, which was increased by ticagrelor and unaffected by either dose of aspirin. In an independent cohort (n = 49, mean age 44, 24 female) we validated that ticagrelor exposure (beta = 0.48, SE = 0.08, p < 0.0001) increases TES scores. In combined analyses of discovery and validation cohorts, when TES levels were calculated using baseline platelet RNA, higher TES levels were associated with lower levels of baseline platelet function (meta-analysis beta = -0.60, standard error [SE] 0.29, P = 0.04) and self-reported bleeding during ticagrelor exposure (meta-analysis beta = 0.28, standard error [SE] = 0.14, P = 0.04). In contrast, we found no associations between bleeding with baseline platelet count, platelet volume, or platelet function.

**Conclusions and Relevance:** Ticagrelor exposure reproducibly and specifically changes a set of platelet transcripts, the baseline levels of which are a biomarker for platelet function and bleeding tendency on ticagrelor.

**Key Points:** *Question:* What are the global effects of ticagrelor exposure on platelets beyond platelet inhibition?

*Findings:* In an experimental human study of different antiplatelet therapies, we comprehensively characterized the effects of ticagrelor on platelet messenger RNA (mRNA). We found that 4 weeks of 90mg twice daily ticagrelor therapy specifically and reproducibly changes the levels of selected platelet mRNA. *At baseline*, volunteers with levels of platelet gene expression that mimic ticagrelor exposure had lower levels of platelet function and when exposed to ticagrelor a greater tendency for minor bleeding.

*Meaning:* By using ticagrelor exposure as a molecular probe, we identified a platelet RNA biomarker that may identify patients at higher risk for ticagrelor-associated bleeding.

## Introduction

Platelet mediated atherothrombotic events such as myocardial infarction (MI) and stroke are major contributors to global morbidity and mortality. Platelet P2Y12 inhibitors, when added to aspirin, are effective in preventing events in high-risk patients, though are associated with an increased risk of bleeding. Ticagrelor is a non-thienopyridine, platelet P2Y12 inhibitor that when added to aspirin prevents MI and stroke in patients with prior^1, 2^ MI. Although effective, potent P2Y12 inhibition with ticagrelor is also associated with major bleeding complications as well as minor (“nuisance” or BARC 1^3^) bleeding that can impact long-term adherence to P2Y12 inhibitor therapy.^4^

Although well known as a potent platelet P2Y12 inhibitor, ticagrelor also has pleiotropic effects. For example, ticagrelor inhibits ENT1, a transporter responsible for the uptake of adenosine from plasma into erythrocytes that subsequently increase systemic, including platelet exposure to adenosine.^5^ In addition, ticagrelor exposure is associated with increased platelet count^6^ -- an effect not ascribed to platelet P2Y12 inhibition^7, 8^. The objective of this study was to use ticagrelor as a molecular probe to comprehensively characterize the on- and off-target effects of ticagrelor on circulating platelets. We tested the hypothesis that ticagrelor associated changes in platelet mRNA underlie variability in platelet biomarkers before and in response to ticagrelor exposure as well as self-reported bleeding.

## Methods

### Overview of clinical studies

We conducted two prospective, previously described (Figure 1A, Friede et al, manuscript under review), independent studies in healthy volunteers conducted in collaboration with the Duke Early Phase Research Unit (DEPRU). Briefly, we conducted a discovery study where HV were randomly exposed to low- or high-dose aspirin, crossover to the alternate aspirin dose, aspirin washout and ticagrelor exposure. Each exposure period lasted four weeks and purified platelets mRNA sequencing and platelet function analysis was performed at each time point, 3 hours after a witnessed ticagrelor dose. The results of the aspirin phase of the analysis are reported separately^9^. Separately, we recruited an additional validation cohort of HV consisting of only a 4 week ticagrelor exposure with identical platelet RNA purification and platelet function analyses.

**Figure 1.**
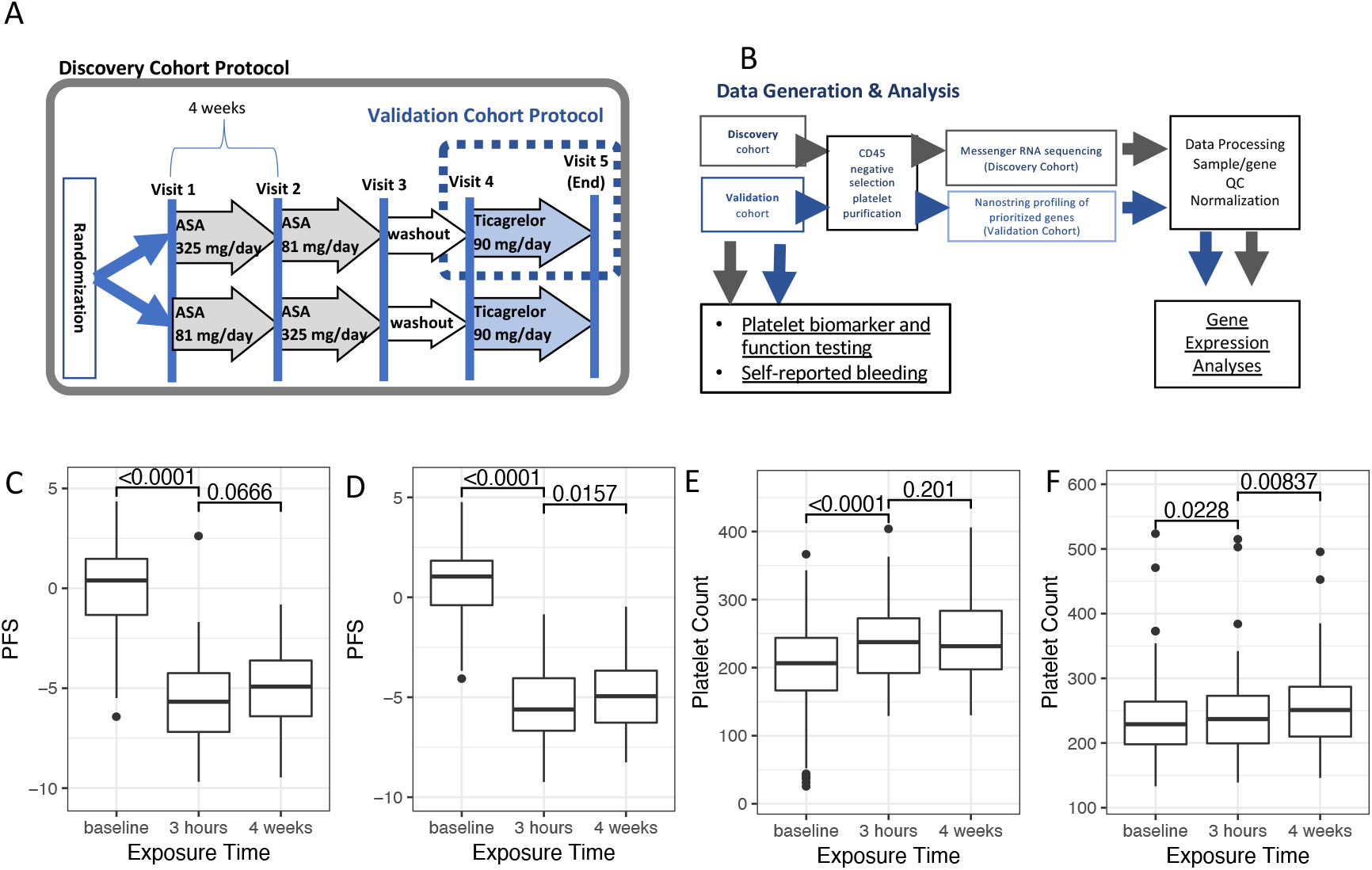
Overview of experimental design and effects of ticagrelor. Panel A: Overview of exposures in discovery and validation cohorts. The discovery cohort began with a baseline visit (Visit 1), randomization to either low- or high-dose aspirin for 4 weeks each (Visit 2), cross over to the other aspirin dose for 4 weeks (Visit 3), aspirin washout (Visit 4), and 4 weeks of ticagrelor exposure (Visit 5). The validation cohort completed a baseline visit (Visit 4) and a single follow up visit after 4 weeks of daily ticagrelor (Visit 5). Panel B: Overview of main outcomes/measures in each cohort. An aggregate measure of platelet function (platelet function score [PFS], y-axis) at baseline, 3 hours after witnessed, 180mg ticagrelor loading dose, and after 3 hours after witnessed final 90mg dose of ticagrelor in discovery cohort (Panel C) and validation cohort (Panel D), with t-test p-values for difference in mean by exposure time denoted at the top. Platelet count at each timepoint in discovery cohort (Panel E) and validation cohort (Panel F), with t-test p-values for difference in mean by exposure time denoted at the top.

Inclusion criteria for both cohorts were age ≥ 30 and ≤ 75 and self-reported non-smokers. Key exclusion criteria were diagnosed bleeding disorder, prior gastrointestinal bleeding, prior gastric/duodenal ulcer, intracranial bleeding, regular use of CYP3A inhibitors or inducers, severe hepatic impairment, current regular use of antiplatelet agents, nonsteroidal anti-inflammatory agents, oral steroids, or anticoagulants, and prescribed daily medications (except oral contraceptives).

Compliance with study drug for the 1st and last dose of each treatment was ensured through a witnessed dose. Between study visits, compliance was monitored in two ways: 1) participants were asked to record the time/date of each drug administration and 2) unused study drug was returned and counted.

Baseline demographic, comorbidity, and concomitant medication data was gathered for each participant. At each visit, phlebotomy for platelet function testing, purified platelet RNA collection, and peripheral blood cell counts were performed using sodium citrate tubes.

Self-reported adverse events were collected at each visit. Treatment specific adverse events related to ticagrelor included: shortness of breath, bleeding (hematoma, nosebleed, gastrointestinal, subcutaneous or dermal), and allergic skin reactions (rash or itching).

The Duke University Institutional Review Board approved all study protocols.

### Platelet function testing and calculations of a platelet function score

Platelet function testing for light transmittance aggregometry and PFA100 at each time point was performed in the Duke Hemostasis and Thrombosis Core Laboratory as previously described.^10^ Briefly, PRP was tested for LTA using epinephrine (0.5, 1, and 10 uM, 12 minutes), ADP (1, 5, and 10 uM, 6 minutes), and collagen (2 and 5 mg/ml, 6 minutes) and the area under the aggregation curve (AUC) recorded. In addition, the PFA100 closure time was measured using a collagen/epinephrine cartridge.

We have previously described^10^ the development and validation of an approach using principal components analysis to summarize the behavior of this panel of platelet function assays measured before and after aspirin exposure. Because the current study utilized two different antiplatelet agents (aspirin and ticagrelor) with different effects on platelet function a modification of our prior approach was required. Instead of performing PCA on the entire dataset (i.e. all time points), we used PCA of visit 1 in the discovery cohort (untreated baseline) samples, standardized the platelet function measure to mean =0, standard deviation = 1, and defined a platelet function score (PFS) as the 1st principal component which explained 47% of the variability in baseline platelet function. The data scaling parameters and loadings matrix were retained for calculating the PFS in additional samples. To generate a PFS in the remaining samples, we standardized the platelet function data as done in the initial calculation and multiplied the standardized data by the visit 1 loadings matrix to generate the on-treatment, aspirin washout, and validation cohort PFS scores.

### Platelet RNA purification

Platelet isolation from human blood using a previously described^11^ leukocyte depletion (LD) procedure is briefly described here. Fresh whole blood (∼40ml) was collected into 4.5ml sodium citrate tubes [3.2% sodium citrate BD vacutainers, VWR], and kept at ambient temperature for processing which was initiated within one hour of phlebotomy. A platelet-rich plasma [PRP] fraction was isolated by slow centrifugation [200 x g for 10 minutes, lowest brake setting]. PRP was transferred into two 15ml conical centrifuge tubes and supplemented with 4 ul 0.5M EDTA per 1ml PRP. The platelets were pelleted from PRP at 1000 x g for 10 minutes resuspended in 2ml sterile filtered Cold Beads Buffer [1X PBS, 0.5% BSA, 2.5mM EDTA]. Human CD45-labeled microbeads [Miltenyi Biotech # 130-045-801] were added to the cell suspension [4ul per 1ml PRP], and incubated for 15 minutes on ice. The suspension was then passed through a magnetic column [MACS Separation LD column# 130-042-901]. The collected platelets were pelleted by centrifugation at 2000 x g for 8 minutes and lysed in 1 ml TRIzol reagent [Life Technologies # 15596018] at room temperature for 10 minutes. Lysates were then transferred to cryovials and stored at minus 80 degrees until RNA extraction.

### RNA isolation

Platelet RNA samples from the same participant were batched together at every stage from platelet RNA extraction through sequencing in order to minimize introducing technical artifacts between visits.

RNA Isolation from purified platelets using a phenol-chloroform method. Platelet lysates thawed on wet ice. Molecular grade Chloroform [200ul m, EMD #CZ1057-6] was added to each 1 ml sample, shaken vigorously by hand for 15 seconds, and allowed to rest for 3 minutes at ambient temperature before centrifugation at 12,000 x g for 10 minutes at 4 degrees. The upper aqueous phase, 500ul 100% isopropanol [Sigma #19516], and 1ul glycogen [Invitrogen #10814-010], were mixed and incubated at ambient temperature for 10 minutes and centrifuged [12,000 x g for 10 minutes at 4 degrees]. After removing the supernatant, the pellet was resuspended with 1ml 75% ethanol [Sigma E-7023], pelleted [8000 x g for 8 minutes at 4 degrees], and eluted in 20ul molecular grade nuclease-free water. DNase treatment was with 2ul (10% volume) 10X DNase Buffer, and 1ul DNase [Invitrogen DNA-free kit #AM1907] for 25 minutes at 37 degrees C. The purified RNA was then precipitated by adding 11.5ul 5M ammonium acetate [USB Ultrapure# 75901], 103.5ul 100% isopropanol and 1ul glycogen at -20 degrees overnight.

### RNA quality analysis and sequencing

Total RNA quality and quantity was assessed using the Agilent Bionalyzer 2100 and the Qubit fluorometer, respectively. Samples with sufficient quantity and quality RNA went on to poly(A) mRNA capture and construction of stranded mRNA-seq libraries from total RNA was achieved using the commercially available KAPA Stranded mRNA-Seq Kit (catalog #KK8421). The manufacturer’s protocol was followed. In brief, mRNA transcripts were first captured from 100ng-200ng of total RNA using magnetic oligo-dT beads. Captured mRNA transcripts were then fragmented using heat and magnesium and reverse transcribed to produce dscDNA. Illumina sequencing adapters were then ligated to the dscDNA fragments and amplified to produce the final RNA-seq library. Libraries were indexed using molecular indexes allowing for multiple libraries to be pooled and sequenced on the same sequencing lane on a HiSeq 4000 Illumina sequencing platform. Each lane consisted of 8-10 samples and yielded about 43 million single-end 50bp sequences per sample.

### Platelet mRNA sequencing alignment and expression quantification

Platelet mRNA sequencing was performed from all available samples and timepoints in the discovery cohort in the Duke Genomics and Computational Biology Sequencing Core Facility. RNA-seq data for mRNA capture samples were downloaded as FASTQ files and processed following as previously described^12^. Briefly, raw single-end sequencing reads were first trimmed to remove adapter sequences and low-quality bases using a sliding window approach with Trimmomatic v0.32 (SE, ILLUMINACLIP:adapters.fasta:2:20:7:1:true, LEADING:5, TRAILING:5, SLIDINGWINDOW:4:10, MINLEN:21). Sequencing quality metrics were determined for both raw and trimmed FASTQ files using FastQC v0.11.4 (Andrews, 2010). Next, reads were aligned with Bowtie2 v2.1.0 (--local, -U) against a custom ribosomal RNA (rRNA) database created by searching the NCBI non-redundant nucleotide database (NR) for Homo Sapiens ribosomal rRNA sequences. To minimize potential rRNA contamination, only unmapped reads which did not align to the rRNA database were then aligned with TopHat2 v2.0.9 to the hg19 transcriptome (Ensemble v74). Expression quantification was done using Cuffquant v2.2.0 and Cuffnorm v2.2.0 (cuffquant –u –b). trimming, alignment, quantification

Cuffnorm gene counts were de-normalized by sample-level internal scale factors to facilitate analysis with RNA-seq count-based protocols implemented in the R package limma. Genes with count < 10 in 80% or more samples were excluded (N = 43423) and samples with ribosomal content > 35% of total reads or that had mean pairwise rank correlations 3 standard deviations lower than the overall mean sample-sample pairwise correlation were excluded (N = 12). For exploratory data analysis, the expression data was normalized using log2 transformed counts per million reads mapped (CPM). For differential expression analysis, expression data was normalized first using trimmed mean of M-values method, then with voom.^13^

### Nanostring analysis

Targeted gene expression of selected transcripts identified using RNAseq was performed in the validation cohort using Nanostring technology.^14^ Platelet housekeeping genes (n = 30) were selected based on their average expression, variability, and compatibility with the Nanostring assay. We selected the 5 genes with the lowest quartile coefficient of dispersion for each quintile of mean expression in the RNAseq that did not cross react with other genes in the Nanostring assay as potential housekeeping genes (Supplemental Table XX). Due to the high expression of *B2M* we used a custom oligo (CAGGCCAGAAAGAGAGAGTAGCGCGAGCACAGCTAAGGC) to attenuate its expression by 95%. In addition to using the manufactured negative controls, we utilized water blanks to generate assay specific background levels. For the majority of samples, 70ng of total RNA was used in each reaction. Nanostring profiling of the mRNA samples was processed using the R bioconductor package Nanostring QC Pro (v1.8). Digital counts were standardized using positive control probe-based scaling factors, background hybridization levels for each probe and lane were established using both negative control probes and water-only samples, and samples were normalized using platelet-specific housekeeping genes (N = 7) for which the mean count was greater than the mean counts of the negative control probes (mean log2 counts > 5) and expression was stable (quartile coefficient of dispersion < 0.2) in the Nanostring expression data.

### Statistical analyses

#### Discovery cohort

Exploratory data analysis of principal components (PC) of the RNAseq data was used to identify any potential technical (e.g. RNA quality/quantity or sequencing related data) or biological (e.g. sex, race, age) between-subjects factors to consider in downstream analysis. These analyses revealed that the concentration of platelet RNA, sex, and sequencing flow cell explained a significant proportion of the variation in expression levels. In addition, we looked for any carryover effects of aspirin by comparing individual transcripts or PCs from the baseline visit vs. the post-aspirin washout visit and found no evidence for significant carryover effect (data not shown). We have also previously found^15^ that platelet gene expression levels are highly correlated within an individual between Visits 1 and Visit 4. Therefore in all subsequent analyses, these two visits were treated equivalently as no-drug treatment visits. We evaluated for levels of leukocyte contamination in the RNAseq data using the CD45/ITGA2B ratio which was lowest in our dataset compared to other published datasets of platelet mRNA seq data (Supplemental Figure 1). Because the level of leukocyte contamination can greatly influence differential expression, we further assessed if CD45/ITGA2B expression was associated with ticagrelor exposure, and found none (p = 0.42). Therefore, no adjustments for the level of leukocyte contamination were made in the primary analysis.

Gene expression was modeled as a function of treatment drug, controlling for sex, starting RNA concentration, flow cell, and repeated measures from individual participants. Repeated measures from the same patient were adjusted for by using subject id as a blocking variable and estimating intra-patient correlation and incorporating this into the model covariance matrix. The linear model fit using the empirical bayes method. Generalized linear hypothesis testing (i.e. contrasts) was used to test differences between specific treatment drugs or combinations of treatment drugs. A false discovery rate of less than 5% was used to determine statistical significance. Analyses were conducted in the statistical program R using packages limma and edgeR.

To further prioritize the list of differentially expressed genes to a manageable number amenable for validation we used three criteria. Genes that met all three criteria were selected for validation.

1. Genes that had average expression (log2 CMP) > 2 in at least one of the treatment groups (untreated or post treated).
2. Differential expression analysis using three alternative quantification and/or normalization starting points (gene level RPKM, transcript-level counts, and transcript-level RPKM) was used to identify robust gene associations. Transcript-level counts were analyzed in the same way as the gene-level counts, described above. RPKM for gene- and transcript-level quantification was log2 transformed and used as input to the linear model, fit using the empirical bayes method, and using generalized linear hypothesis testing to test differences between specific treatment drugs or combinations of drugs. Differentially expressed genes with additional evidence of treatment response in any of these three analyses were prioritized for replication.
3. To fulfill the third criterion, one of the following had to be met:

a. Regularized logistic regression (lasso) was used to construct a cross-validated, multi-gene model of drug exposure and the differentially expressed genes in this model were prioritized for replication. Briefly, participants were split 80%/20% for training and testing datasets. Within the training data set, participants were partitioned into 10-folds for cross validation, ensuring all samples from a participant fall within a single fold. Within each cross validation, genes were filtered by differential expression re-analysis, with genes having an adjusted p-value < 0.05 as input to the lasso model. The resulting models were preserved from each cross validation and genes were summarized by the number of cross validation models in which they appeared with non-0 weights. Differentially expressed genes appearing in any of the models were prioritized for replication.
b. Weighted gene co-expression network analysis (WGCNA) -- We applied WGCNA to the entire platelet mRNA sequencing data (i.e. 14333 genes in 312 samples). Using the default WGCNA parameters, we identified 111 networks (or “modules”). To further reduce the number of modules, we used hierarchical clustering of the aggregate expression of each module to merge modules with similar aggregate gene expression (Supplemental Figure XX). The result of this process was the identification of 30 unique modules that each represent the aggregate expression of the genes within each module. The attributes of these modules is listed in Supplemental Table 6. The aggregate expression of all genes assigned to a module can be summarized using principal components analysis (PCA) where the 1st principal component (named eigengene) is used as a summary measure of module gene expression. Because each module eigengene can be thought of as the aggregate expression of all of the genes in that module, we can use the eigengene value to test for association with ticagrelor exposure to assess the behavior of the entire module. Each module eigengene was tested for association with ticagrelor using an FDR < 5% cutoff. To select genes within each module, we selected the top 10 genes within each differentially expressed module that were themselves differentially expressed by ticagrelor at FDR < 5% (i.e. part of the original 3402) and 1) their connectivity to the module or 2) their correlation with each module eigengene.

#### Validation cohort

Differential expression was tested using linear mixed effect models, modeling log2 normalized gene expression as a function of ticagrelor treatment and sex, with a participant-level random intercept. Significance of the treatment effect was assessed using a likelihood ratio test of nested models. The ticagrelor expression signature (TES) was calculated as the mean difference of up regulated genes and down regulated genes (directionally based on the mRNA-seq discovery analysis). The TES was tested for association with treatment using the same mixed effect model as used for the individual genes. The linear relationship between the platelet function score and the TES was assessed using 3 models - 1) a linear mixed effect model using both on and off treatment samples while controlling for sex, treatment, and participant level repeated measures, 2) a pre-treatment linear fixed effect model with sex as covariate, and 3) a post-treatment linear fixed effect model, also adjusting for sex.

#### Meta-analyses

Parameter estimates from the TES associations with platelet function measures, platelet count, and bleeding events were combined across the discovery and validation cohorts using meta-analysis. Parameter estimates were weighted using the inverse variance weights and summed to generated the combined estimates and standard errors, using the R package meta. Both the fixed effect and random effect model for between study variances were used. Significance was assessed using standard normal assumptions for the Z score derived from the combined estimates and standard errors.

## Results

### Study overview and participant characteristics

A summary of the experimental protocol and data analysis for the Discovery and Validation cohort studies is summarized in Figure 1A and 1B, respectively. The numbers of patients and their baseline characteristics are in Supplemental Tables 1 and 2, respectively. In the discovery cohort, healthy volunteers (HV) had a mean age of 43 years, were 67% female, 44% white, and 43% black. In the validation cohort, HV had mean age of 43 years and were 58% female, 55% white, and 33% black.

### Platelet biomarker outcomes

As expected, an 180 mg ticagrelor loading dose significantly reduced platelet function as measured using an aggregate all platelet function assays, a platelet function score (PFS, Figures 1C and 1D), as well as each individual assay of platelet function (Supplemental Table 3 and 4) in both cohorts. While there was no change in mean platelet volume (MPV) with ticagrelor, platelet counts increased with a single loading dose of ticagrelor (Figure 1E and 1F).

After 4 weeks of twice daily dosing with 90 mg ticagrelor, platelet function remained inhibited. However, compared to the PFS measured 3 hours after the initial ticagrelor loading dose, we observed a rebound in platelet function 3 hours after the final ticagrelor maintenance dose in the PFS (mean change in PFS between 4 weeks and 3 hours in validation cohort = 0.5, standard error [SE] = 0.2, p = 0.02, Figure 1C). This increase in the PFS towards baseline levels of platelet function was of similar magnitude in the discovery cohort (mean change = 0.45, SE = 0.28, p = 0.1, Figure 1D). In a combined analysis of discovery and validation cohorts, the change in PFS between 3h and 4 weeks was significant (inverse variance weighted meta-analysis effect = 0.49, 95% confidence interval: 0.15-0.82, p-value = 0.004) and was driven by trends towards higher ADP and epinephrine induced aggregation at 4 weeks vs. 3h (Supplemental Tables 3 and 4). Platelet counts continued to modestly increase with daily ticagrelor dosing (Figure 1E and 1F) and MPV remained stable (Supplemental Tables 3 and 4).

Therefore, an initial dose of ticagrelor inhibits platelet function and lowers platelet count. During daily ticagrelor dosing, platelet function increases towards baseline levels, while platelet count continues to rise.

### Effect of ticagrelor exposure on global platelet gene expression

After quality control analysis of the platelet mRNA sequencing data, there were 14333 transcripts available for statistical analysis in 312 samples (baseline, post-aspirin, aspirin washout, and post-ticagrelor). Using a false discovery rate (FDR) cut off of 5% to define statistical significance, ticagrelor exposure was associated with the upregulation of 1820 genes and the down regulation of 1589 genes (Figure 2A). We performed sensitivity analyses to explore the potential confounding effects of 1) including a measure of residual leukocyte contamination (CD45/ITGA2B ratio) 2) including platelet count measured in whole blood or 3) excluding platelet RNA concentration after leukocyte depletion as covariates. In each sensitivity analysis we found that 3381 (99%%), 2720 (80%), and 3075 (90%) of the 3409 differentially expressed genes were retained. While the inclusion of CD45/ITGA2B or platelet count identified small numbers of differentially expressed genes (296 and 86, respectively) that were not originally identified, the exclusion of platelet RNA concentration resulted in an additional 3290 differentially expressed genes for which the association with ticagrelor exposure was confounded by differences in RNA concentration.

**Figure 2.**
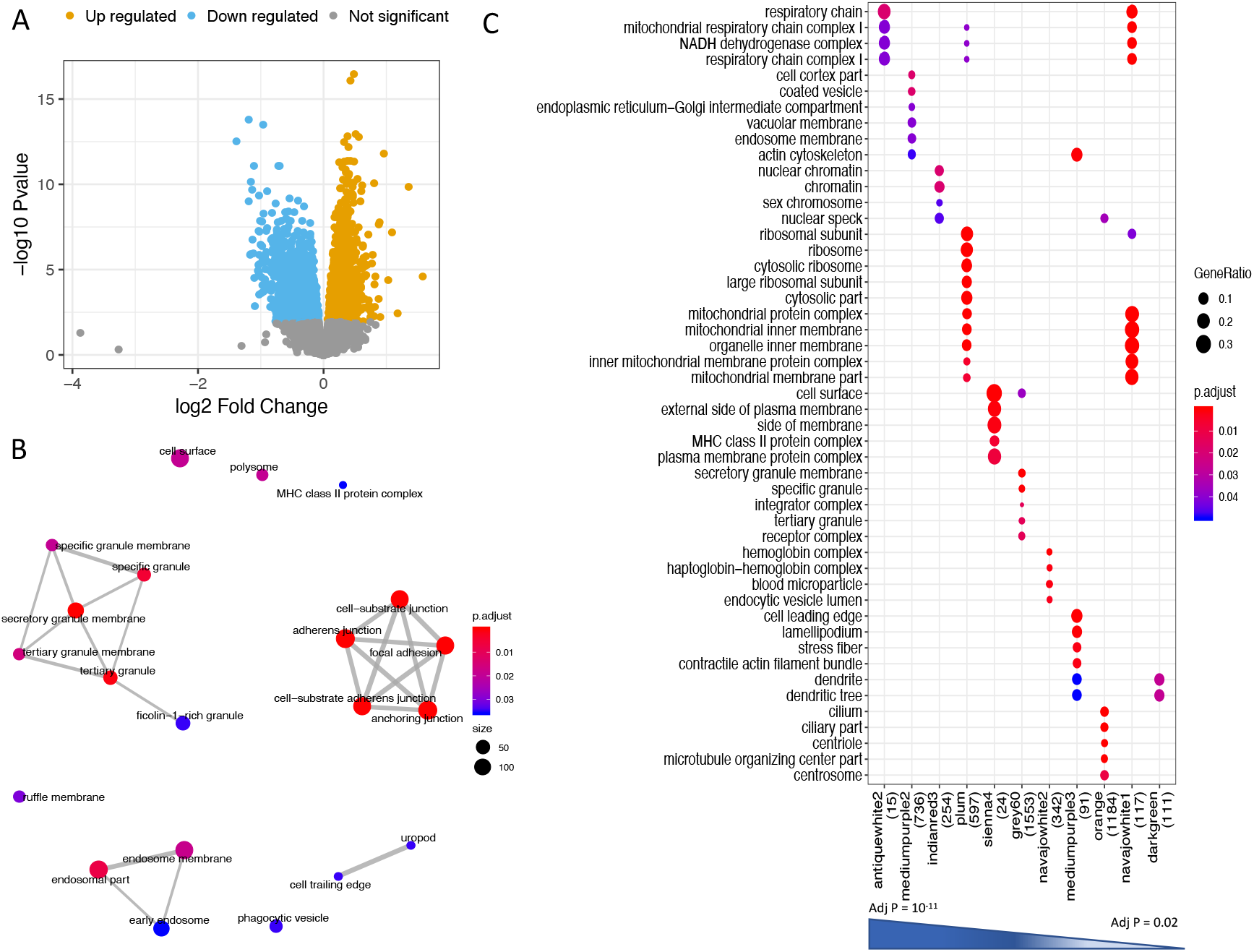
Pathways analysis of differentially expressed genes by ticagrelor. Panel A: Volcano plot of 14333 platelet transcripts tested for differential expression with ticagrelor. The log2 fold change (lx-axis) for each gene is plotted against the –log10 of the p-value (y-axis) for that gene. Colored points represent genes below the false discovery threshold of 5%. Blue points represent down regulated genes while orange colored points represent upregulated genes. Panel B: Gene ontology (GO) analysis of Cellular Components enriched in the 3409 genes associated with ticagrelor exposure. The thickness of the edges connecting nodes are scaled based on the number of genes overlapping the connected pathways divided by the number of unique genes in the union of the connected pathways from the list of differentially expressed genes. Panel C: GO analysis of weighted gene co-expression network modules that change in response to ticagrelor exposure. Column names represent individual modules or networks (each named a different color, number of genes in each module in parentheses) and are listed in descending order of strength of association between the module eigengene value and ticagrelor exposure.

We used two complementary approaches to categorize the functional effects of ticagrelor on platelet gene expression. First, we used Gene Ontology (GO) enrichment analysis of all 3409 differentially expressed genes to identify the following cellular components within the platelet that may be impacted by ticagrelor: 1) secretory granules 2) cell-extracellular matrix junctions and 3) endosomes (Figure 2B). To gain further insight on the biological networks impacted by ticagrelor we used weighted gene co-expression network analysis (WGCNA) to identify sets of co-expressed genes (or “modules” each named a different color) within the RNAseq dataset in an unsupervised manner. Of the 30 unique modules identified, the aggregate expression of 21 was associated (FDR < 5%) with ticagrelor exposure. (Supplemental Table 5). We used GO to annotate each of the differentially expressed modules with known biological pathways and 11 of 21 had significant (adjusted p-value < 0.05) GO annotations (Figure 2C). In addition to identifying modules - and by extension the co-expressed genes underlying each module - annotated to endosomes (“mediumpurple2”), and secretory granules (“grey60”) we additionally identified mitochondrial components (“antiquewhite2”, “plum”, and “navahowhite1” modules) and ribosomes (“plum” and “navahowhite1”) as being affected by ticagrelor exposure. There were 10 modules that were differentially expressed by ticagrelor but could not be annotated into a known GO or KEGG (data not shown) category and therefore their function was not readily apparent.

Therefore, ticagrelor affects specific networks of co-expressed platelet genes that highlight platelet secretory or adhesion functions.

### Validation of Ticagrelor’s effects on platelet gene expression

In order to reduce the set of differentially expressed genes to a small number for validation and at the same time retain the biological effects of ticagrelor on global platelet gene expression, we used a set of prioritization criteria (Figure 3A). After applying these criteria to the list of differentially expressed genes there were 111 genes (Supplemental Table 6) that met all three criteria that we used to define a Ticagrelor Exposure Signature (TES) that we summarized into an aggregate TES score. To confirm that the subset of 111 genes captures similar information present in the full 3402, we correlated the scores using both sets of genes and found that a score based on the subset of 111 genes was strongly correlated (r = 0.92, p <0.001) with one constructed on all 3402 genes (Figure 3B). As expected based on its definition, the TES score increased in response to ticagrelor in the discovery cohort and was unaffected by low-or high-dose aspirin exposure (Figure 3C).

**Figure 3.**
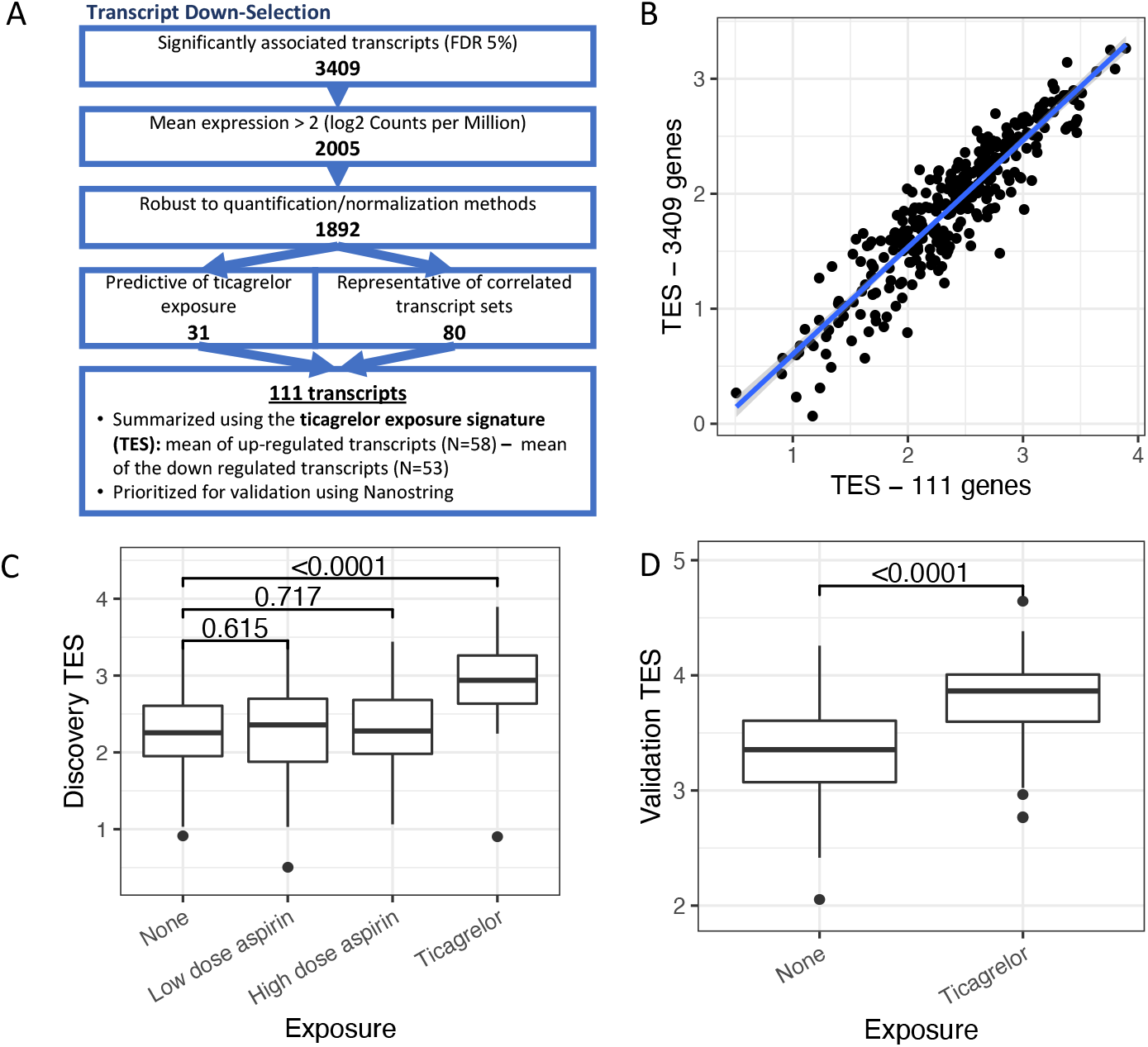
Generation of a ticagrelor exposure signature (TES). Panel A application of prioritization criteria to filter list of 3409 differentially expressed genes to 111 transcripts used to define the TES. Panel B Scatter plot of TES defined by full list of 3409 differentially expressed genes (y-axis) vs. 111 gene that passed prioritization criteria (x-axis). Panel C: Box-whisker plots of TES levels based on 111 genes in Discovery cohort in absence of drug exposure, after low- and high dose aspirin, and ticagrelor exposures. Panel D: TES score based on 84 of 111 prioritized TES genes measured using Nanostring in Validation cohort.

To independently validate ticagrelor’s effects on individual platelet gene expression, we used a custom Nanostring assay to measure the expression of TES genes in the validation cohort (n = 42 that had quality RNA). Because Nanostring is an alternative measure of gene expression compared to mRNAseq, we first refined the TES genes to those that exhibited positive correlation (r > 0.25) with platelet mRNA seq in a subset of discovery cohort samples, which reduced the number of TES genes from 111 to 84 for validation testing. At an individual gene level, 36 of 84 genes (42.9%, FDR < 5%) were associated with ticagrelor exposure in the validation cohort. Furthermore, the direction of ticagrelor’s effect on platelet gene expression was positively correlated for each gene in the discovery vs. validation cohorts (r = 0.77, P = <0.0001).

In order to validate ticagrelor’s effects on TES scores, we calculated a TES score using the 84 filtered genes using the same definition as in the discovery cohort. In the validation cohort, we found that ticagrelor increased TES scores as in the discovery cohort (beta = 0.48, SE = 0.08, p < 0.0001, Figure 3D). In a sensitivity analysis, we constructed TES scores using all 111 genes measured by Nanostring and continued to find association with ticagrelor exposure (beta = 0.45, SE = 0.07, P < 0.0001)

Therefore the TES represents a set of platelet genes that reproducibly and specifically changes in response to ticagrelor exposure but not aspirin.

### Ticagrelor responsive genes and platelet biomarker measurements

To evaluate the extent to which the network of genes affected by ticagrelor exposure correlate with platelet biomarkers we compared the TES score with the composite platelet function score (PFS), platelet count, and MPV, measured at each time point in the combined discovery and validation cohorts. At baseline we observed an association between higher TES and lower PFS (meta-analysis beta = -0.6, 95% confidence interval: [-1.6, -0.04], p = 0.04, Figure 4A). In contrast, after 4 weeks of ticagrelor exposure, TES levels were not associated with on-treatment platelet function (meta analysis p = 0.74). While no individual component of PFS was significantly associated with TES scores, their directions of association were all consistent with the aggregate measure: higher TES scores reflect lower levels of baseline platelet function (Figure 4B). We found no associations between TES levels at baseline or on ticagrelor with MPV or platelet count (meta-analysis p-values > 0.2).

**Figure 4.**
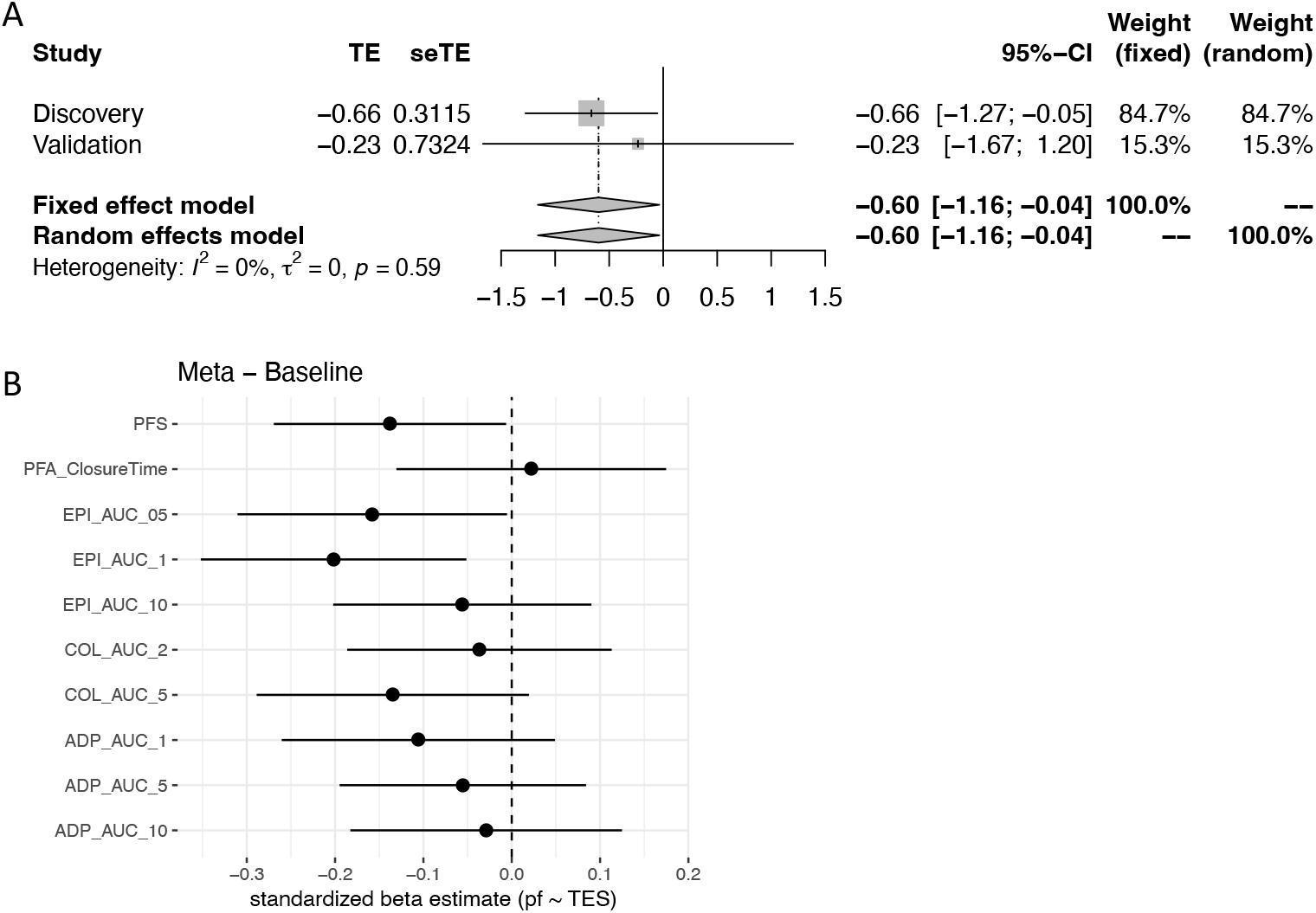
Ticagrelor exposure signature score associations with baseline platelet function measures. Panel A: Forest plot of association between TES scores measured in Discovery and Validation cohort with the aggregate measure of platelet function, platelet function score, and combined using meta-analysis. Panel B: Forest plot of meta-analysis beta estimates for association between TES score and individual measures of baseline platelet function and the aggregate PFS.

Therefore, at baseline, healthy volunteers with naturally occurring, higher platelet TES scores - potentially reflecting a baseline effect on platelets that is similar to the effect of ticagrelor - are characterized by also having lower levels of platelet function.

### Analysis of Self-Reported Bleeding outcomes

Self-reported bleeding was recorded in a small number of participants in the discovery (N = 6) and validation (N = 3) cohorts during ticagrelor exposure. Based on the association of higher TES scores with ticagrelor exposure and decreased platelet function at baseline, we explored associations with self-reported bleeding outcomes. In a combined analysis of both cohort and after adjusting for treatment effects, higher TES scores were associated with bleeding (meta-analysis beta = 0.28, SE = 0.14, p = 0.04, Figure 5A).

**Figure 5.**
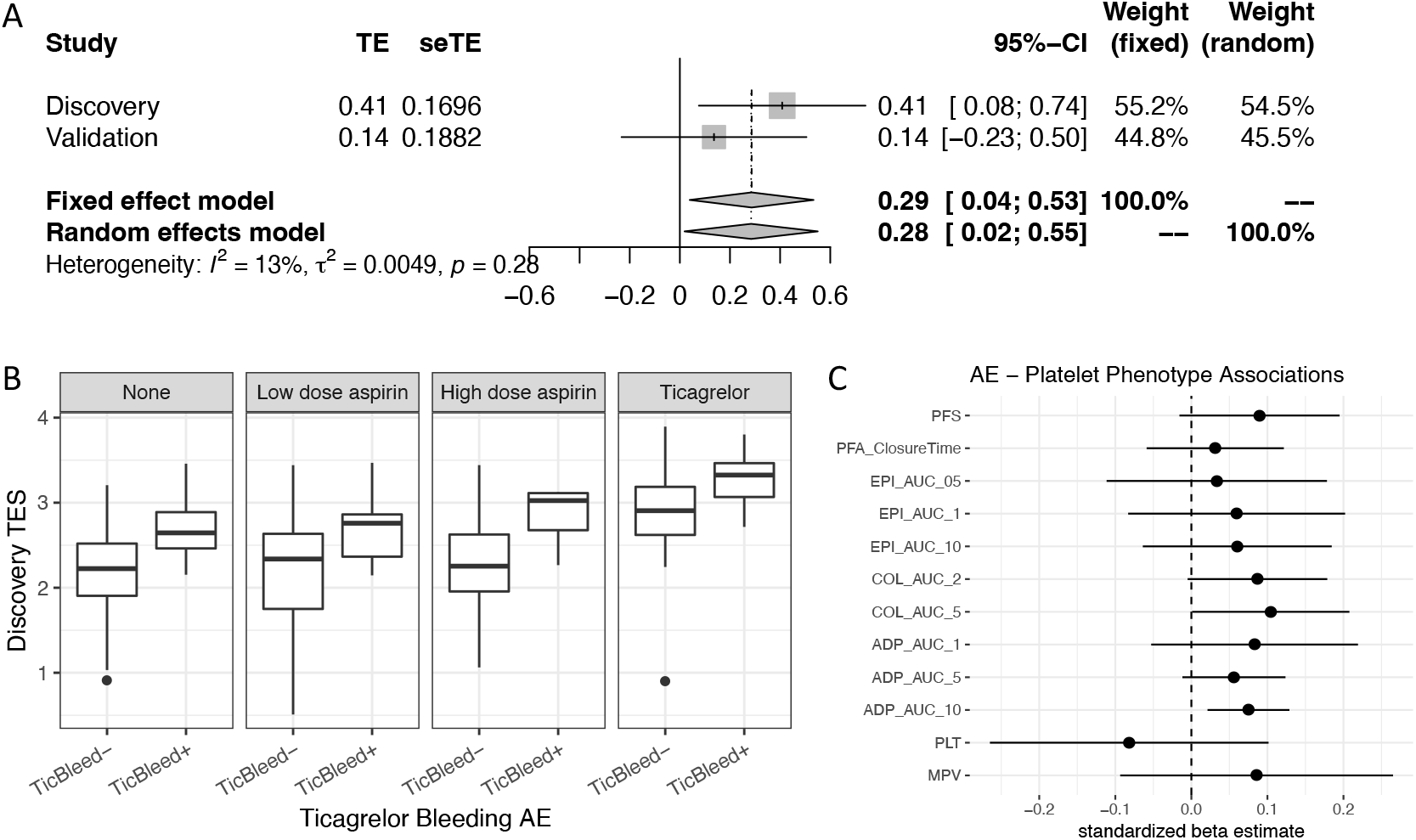
Ticagrelor exposure signature score associations with self-reported bleeding. Panel A: Forest plot of beta estimates of association between TES and self reported bleeding during ticagrelor exposure status adjusted for treatment in Discovery and Validation cohorts and combined through meta-analysis. Panel B: Box-whisker plots of TES scores (y-axis) in Discovery cohort vs. bleeding outcome during ticagrelor exposure stratified by drug exposure. Panel C: Forest plot for beta estimates of platelet biomarker associations with bleeding during ticagrelor exposure from the combined analysis of the Discovery and Validation cohorts.

To further assess the relationship between TES, drug exposure, and bleeding, we stratified TES levels by bleeding and by visit in the discovery cohort and found that patients who reported bleeding during ticagrelor exposure had higher TES levels at baseline that remained during aspirin and ticagrelor exposure compared to non-bleeders (Figure 5B, p = 0.02). There was no difference in the magnitude of change in TES between those who did vs. did not report bleeding (p = 0.4). In contrast to TES levels, none of the baseline platelet function parameters individually, the aggregate PFS, platelet count, or MPV were associated with bleeding in this cohort. (Figure 5C)

Therefore, in addition to having relatively decreased baseline platelet function, patients with higher TES levels are more likely to experience minor bleeding during ticagrelor exposure.

## Discussion

Platelet P2Y12 inhibitors such as ticagrelor are widely used to prevent platelet-mediated thrombotic events such as myocardial infarction and stroke. While the main mechanism of action of ticagrelor is to inhibit platelet P2Y12, it is unknown to what extent these agents exhibit additional, potential off-target effects that may explain heterogeneity in clinical outcomes. We used genome wide profiling of platelet mRNA before and after ticagrelor exposure to comprehensively characterize the platelet response to ticagrelor exposure. We tested the hypothesis that tiacgrelor’s effects on platelet gene expression are associated with platelet biomarker and bleeding outcomes in an experimental drug-exposure model of healthy, human volunteers. We identified a unique set of transcripts that reproducibly change in response to ticagrelor exposure that we summarized as the Ticagrelor Exposure Signature (TES). We found that ticagrelor uniquely upregulates TES levels, that higher TES levels are associated with lower levels of baseline platelet function, and that higher baseline TES levels identify patients prone to minor bleeding during ticagrelor exposure. In summary, through a global transcriptional analysis of ticagrelor’s effects on platelets, we identify novel genes and pathways that contribute to platelet function and may be useful bleeding biomarkers.

Platelets are anucleate cells and therefore are incapable of *de novo* transcription. Instead, platelets are endowed with a repertoire of mRNA during thrombopoiesis as part of a regulated process. Changes in megakaryocyte transcription can be reflected, in part, in the mRNA content of circulating platelets as is seen in the case of mutations that alter transcription factor binding. We do not believe the changes we observed due to an “artificial” differential expression caused by ticagrelor’s effects on platelet counts because we retain a high proportion (> 80%) of transcripts when 1) controlling for platelet RNA concentration (a surrogate for platelet RNA content since each participant contributes a fixed blood volume) in all analyses and 2) platelet count/volume in sensitivity analyses. Our study was not designed to elucidate the mechanism by which ticagrelor alters platelet mRNA levels. However, to explore the extent to which ticagrelor may have effects on transcription, we used the Connectivity Map database using the top differentially expressed platelet genes as an input and identified 60 perturbations (of 8796) that produced similar gene expression changes as ticagrelor, including two related to ticagrelor’s known off-target effects on adenosine turnover: knockdown of *ADK* (adenosine kinase) and *NT5E* (ecto-5’-nucleotidase or CD73). Although ticagrelor was not included in the Connectivity Map library, neither alternative P2Y12 inhibitors (clopidogrel and ticlopidine), nor manipulation of *P2YR12* (knockdown or overexpression) mimicked effects of ticagrelor on gene platelet gene expression in this dataset. In our study we also found that neither low-nor high-dose aspirin affected TES levels. Therefore, we hypothesize that ticagrelor or one of its metabolites may have unique, off-target effects on megakaryocyte transcription that in turn alters the content of circulating platelets. The association with higher TES levels and decreased baseline platelet function suggests that the effect of ticagrelor on platelet mRNA may represent an effect of ticagrelor on inhibiting platelet function that is independent of P2Y12 receptor inhibition.

A small number of participants reported bleeding during the 4-week ticagrelor exposure, which were all classified as “nuisance” bleeding events (easy bruising, epistaxis, and menorrhagia) and mild in severity as they did not require medical attention or cessation of ticagrelor. Analysis of TES levels in patients who bled on ticagrelor found an overall association with higher TES levels compared to non-bleeders (Figure 5A). We found no difference in the magnitude of change in TES levels with ticagrelor exposure between those who did vs. did not bleed on ticagrelor. Instead, we found that participants who bled had higher TES levels at baseline, prior to initiating ticagrelor. Our finding that higher TES levels at baseline are associated with lower platelet function is consistent with prior associations of lower, on-treatment platelet function on clopidogrel and major bleeding.^16^ That we did not find any platelet biomarkers associated with minor bleeding in our study (Figure 5D) suggests that TES genes are reporting on an aspect of platelet physiology that may be relevant for bleeding in addition to *ex vivo* platelet function. For example, GO analysis of genes differentially expressed with ticagrelor identified cell-extracellular matrix junctions which may be important for maintaining hemostasis *in vivo* but not for cell-cell adhesion needed for platelet aggregation assays. Further validation of this association and extension to major bleeding will be important to further validate our findings.

That participants who bled on ticagrelor had higher TES levels during the three timepoints *before* initiating ticagrelor (Figure 5B) suggests that there are important genetic and/or environmental factors that influence TES levels. A cross-reference to the largest platelet expression quantitative trait loci (eQTL) database^17^ to date demonstrated that 150 of the 3409 ticagrelor responsive genes have at least one, significant, cis-eQTL. That ticagrelor exposure results in the highest TES levels in those who bled suggests that there may be a “threshold” above which TES levels are associated with bleeding. Patients with levels above this threshold at baseline may be at higher bleeding risk regardless of the perturbation (antiplatelet or anticoagulant) whereas those with elevated but sub-threshold baseline TES levels may be prone to bleeding on ticagrelor, specifically. Therefore, the TES may represent a novel, baseline bleeding risk marker that is independent of traditional platelet biomarkers and may be a useful companion diagnostic for patients considering initiating antithrombotic therapy.

Despite the strengths of our study: prospective assessment of drug effects in an experimental model, independent validation, robust assessment of gene expression changes there are limitations. First we do not extend our findings to platelet protein; future mechanistic studies focused on the biological effects of ticagrelor on platelet mRNA will require this. Second, our trial did not include a placebo control and all patients in the discovery cohort were first exposed to aspirin. We believe the independent validation in a *de novo* cohort that was only exposed to ticagrelor and the lack of effects during aspirin exposure or aspirin washout in the discovery cohort address this limitation. Third, we acknowledge that clopidogrel is the most commonly prescribed platelet P2Y12 inhibitor and that future work will be needed to understand the extent to which the effects of ticagrelor extend to other platelet P2Y12 inhibitors. Fourth, the associations between TES and bleeding, were exploratory will require additional validation in larger datasets. Last, we do not yet know how TES levels may explain variability in ticagrelor’s efficacy in preventing ischemic endpoints. However, we are unaware of any existing biorepositories with banked platelet RNA and ischemic outcomes during ticagrelor exposure.

In summary, by using ticagrelor as a molecular probe to characterize the response to this antiplatelet agent, we identified a set of platelet genes that change in response to ticagrelor exposure. Patients with an effect in their platelet mRNA at baseline that recapitulates the effect with ticagrelor have lower levels of platelet function and are prone to bleeding during ticagrelor exposure.

## Data Availability

Gene Expression Omnibus (GSE158765)

## Figure legends

**Supplemental Figure 1. CD45/ITGA2B ratios in this and other published platelet RNAseq datasets**. The amount of leukocyte contamination was estimated as the ratio of leukocyte marker CD45 (encoded by *PTPRC*) to platelet gene *ITGA2B*. This in the current study is compared to three other publicly available platelet RNAseq datasets.

**Supplemental Figure 2. Dendrogram of WGCNA modules**. Hierarchical clustering of the aggregate expression of the 111 individual modules was performed. Modules with similar expression are closer to each other along the x-axis and similarities (or dissimilarities) between modules is indicated by the vertical distance between modules. The red-horizontal line depicts the cut-point for merging modules. Genes within modules that are part of a tree that forms below the red line are merged into a single module. For example the genes belonging to the darkolivegreen4 and honeydew1 modules are merged into a new module named darkolivegreen4 (not shown). In contrast, the genes in the darkmagenta and firebrick3 modules remain separate modules with the same names.

**Supplemental Table 1.**
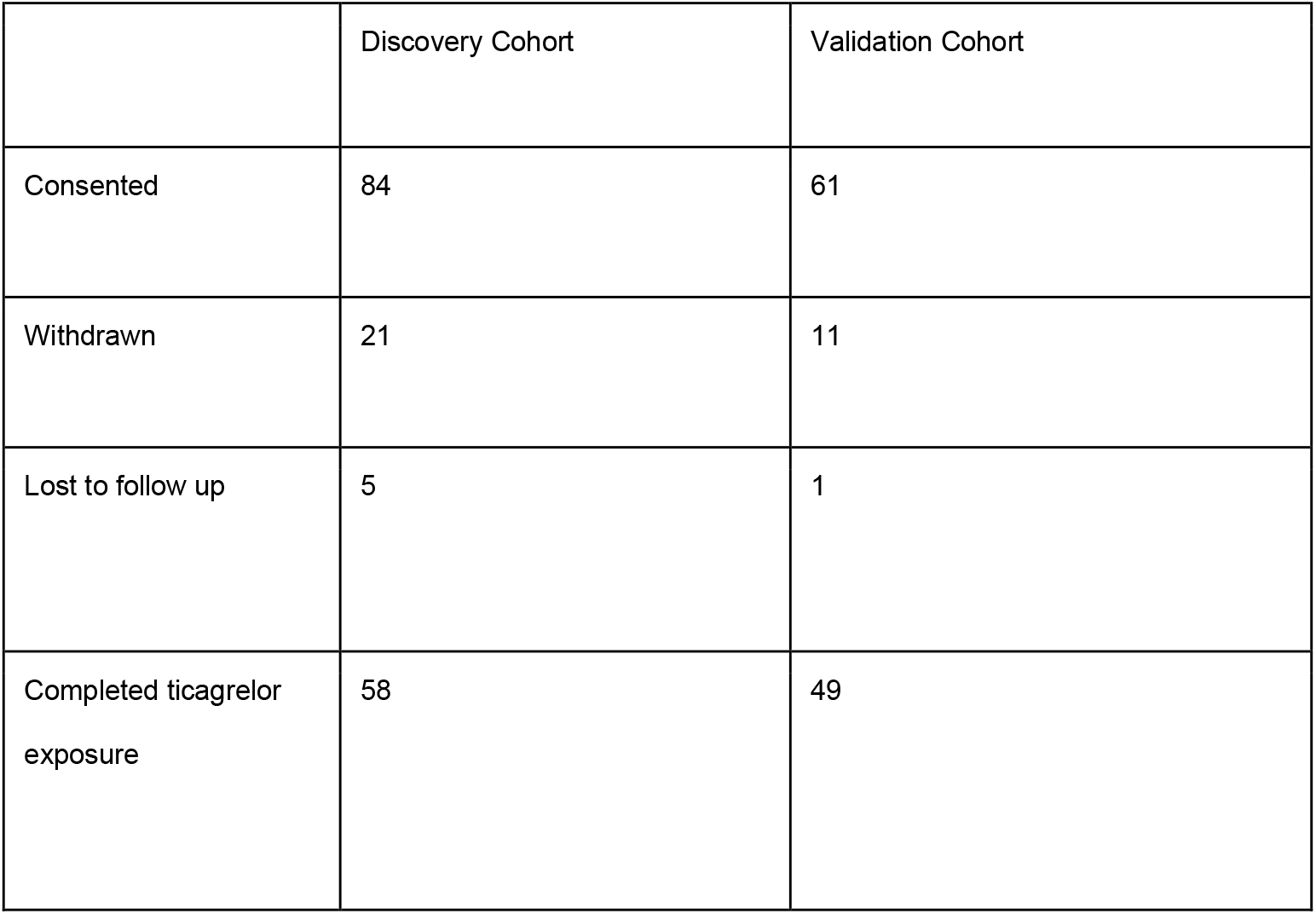
Numbers of participants in each cohort

**Supplemental Table 2.**
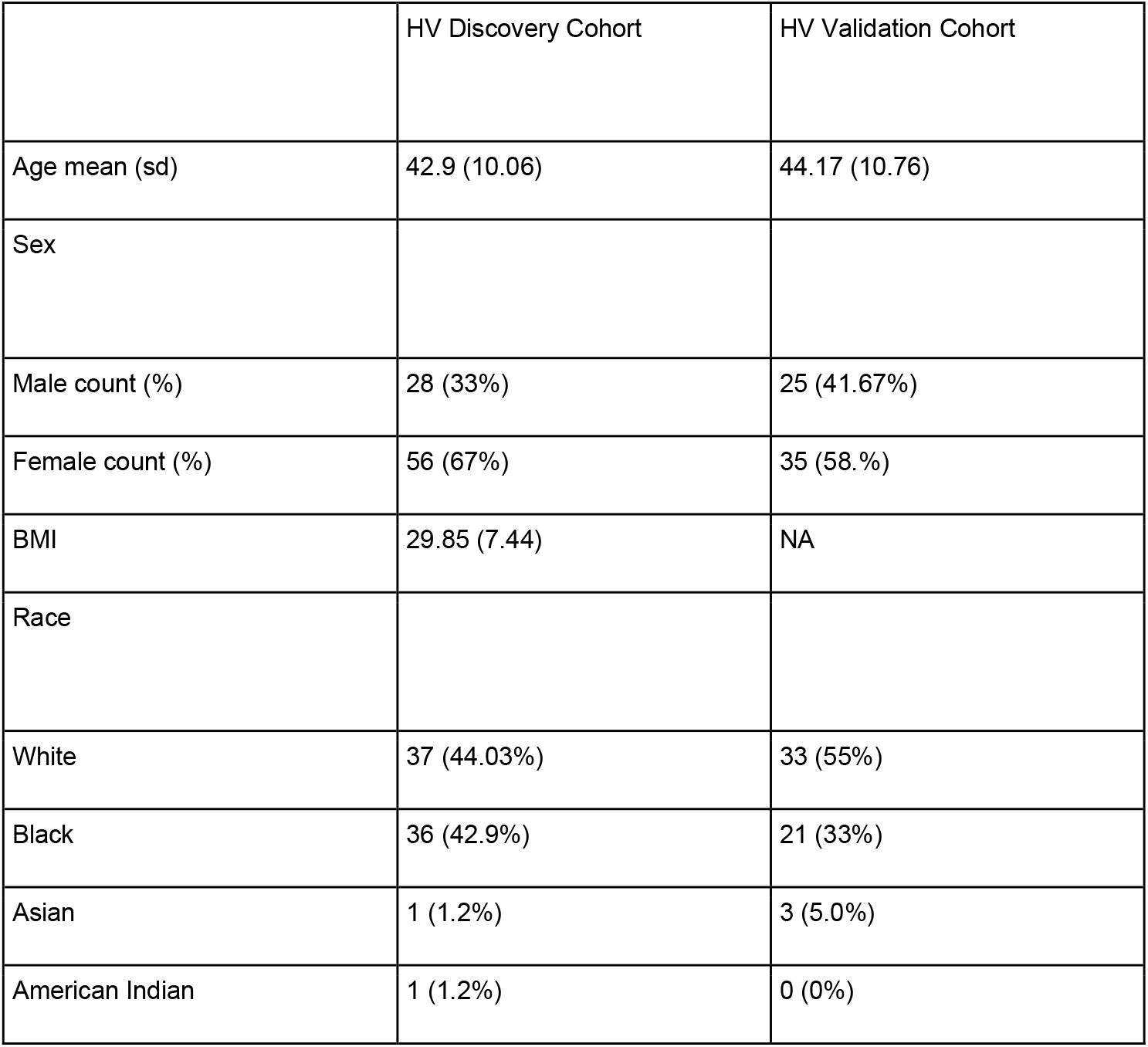
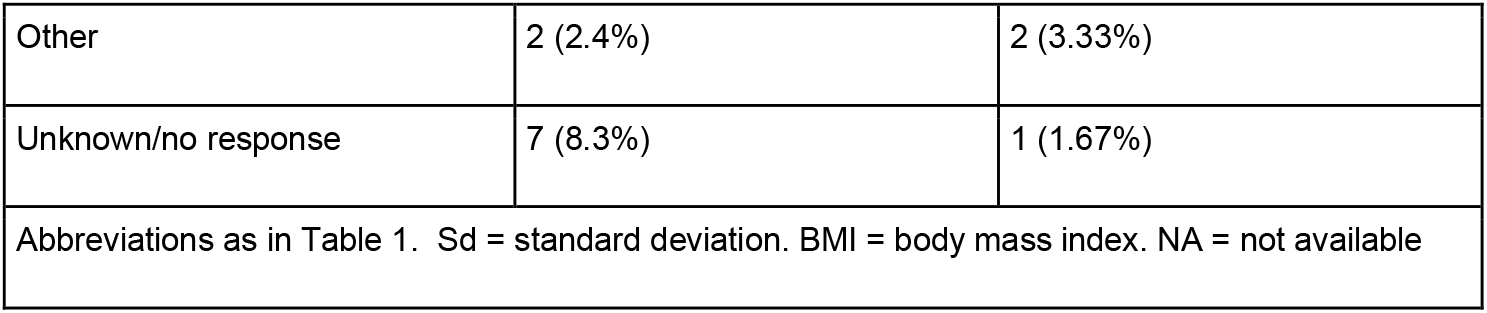
Baseline characteristics participants

|  | HV Discovery Cohort | HV Validation Cohort |
| --- | --- | --- |
| Age mean (sd) | 42.9 (10.06) | 44.17 (10.76) |
| Sex |  |  |
| Male count (%) | 28 (33%) | 25 (41.67%) |
| Female count (%) | 56 (67%) | 35 (58.%) |
| BMI | 29.85 (7.44) | NA |
| Race |  |  |
| White | 37 (44.03%) | 33 (55%) |
| Black | 36 (42.9%) | 21 (33%) |
| Asian | 1 (1.2%) | 3 (5.0%) |
| American Indian | 1 (1.2%) | 0 (0%) |
| Other | 2 (2.4%) | 2 (3.33%) |
| Unknown/no response | 7 (8.3%) | 1 (1.67%) |

Abbreviations as in Table 1. Sd = standard deviation. BMI = body mass index. NA = not available

**Supplemental Table 3.**
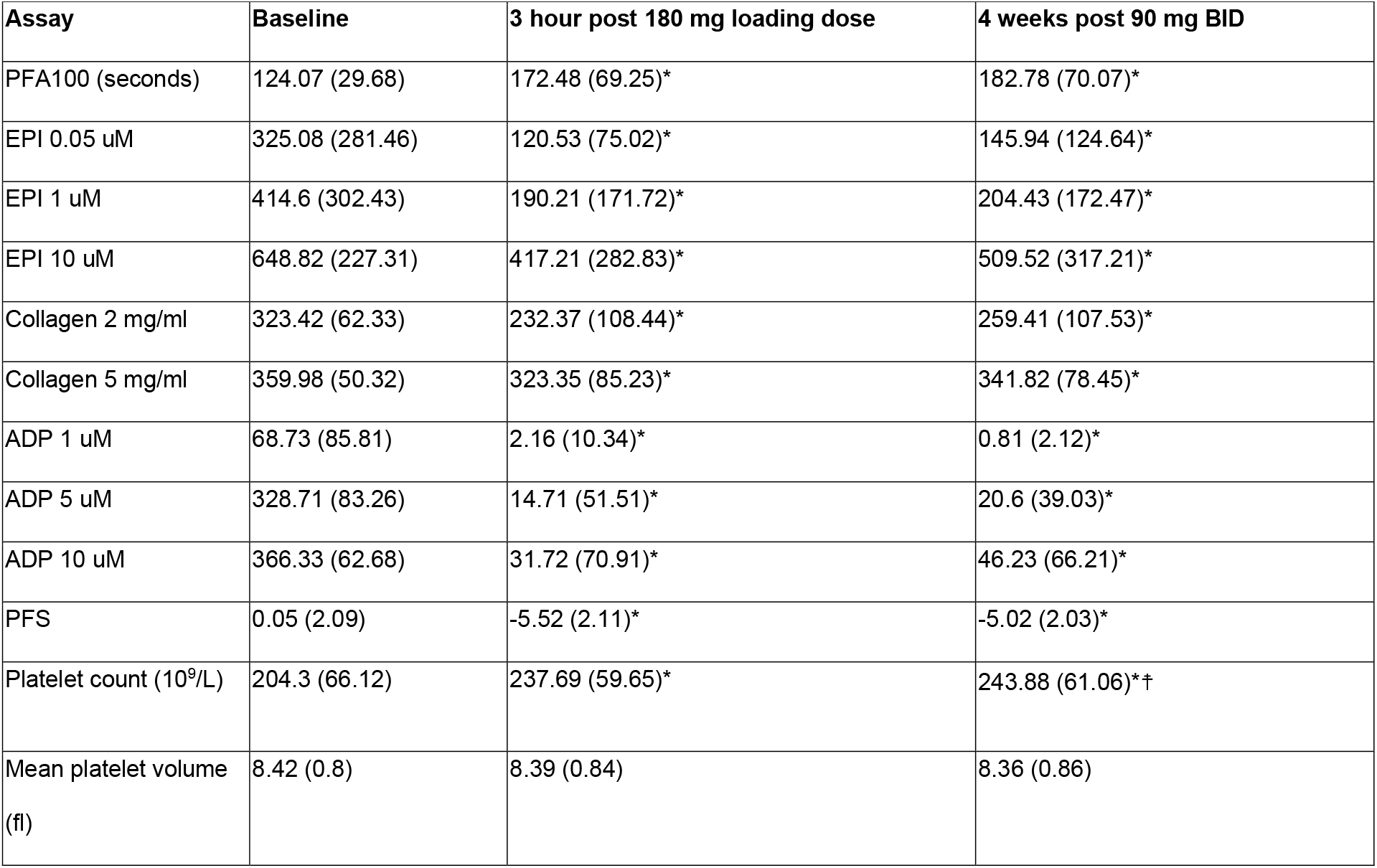
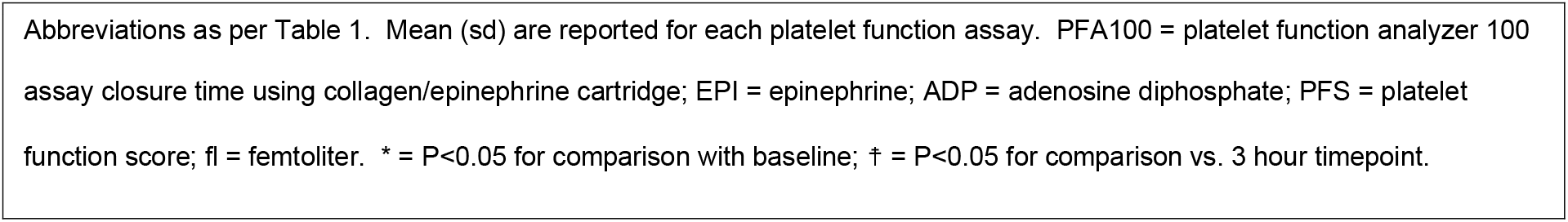
Platelet function test results in discovery cohort

**Supplemental Table 4.**
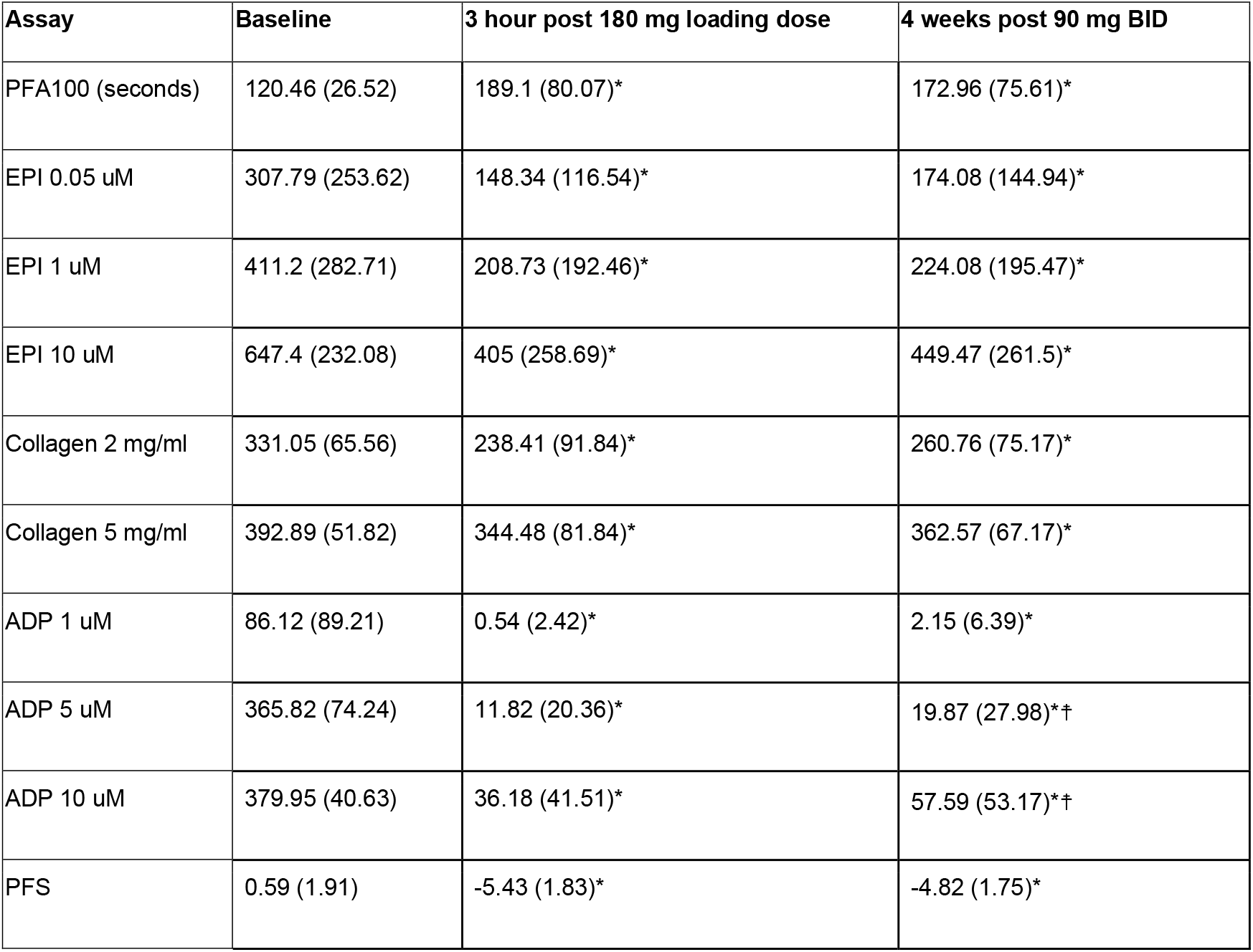
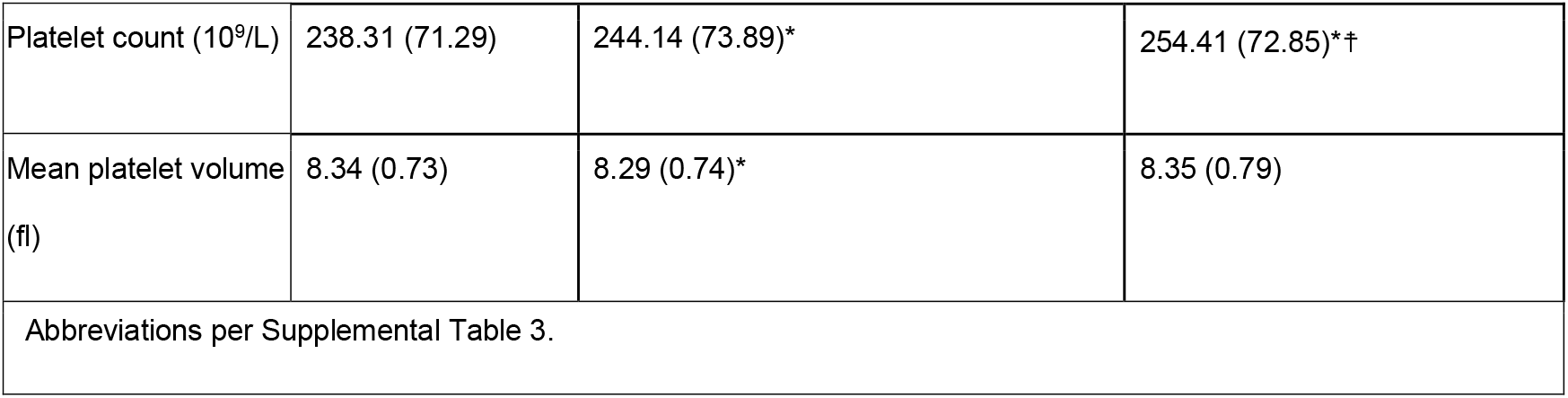
Platelet function in Validation cohort

| Supplemental Table 4. Platelet function in Validation cohort |  |  |  |
| --- | --- | --- | --- |
| Assay | Baseline | 3 hour post 180 mg loading dose | 4 weeks post 90 mg BID |
| PFA100 (seconds) | 120.46 (26.52) | 189.1 (80.07)* | 172.96 (75.61)* |
| EPI 0.05 uM | 307.79 (253.62) | 148.34 (116.54)* | 174.08 (144.94)* |
| EPI 1 uM | 411.2 (282.71) | 208.73 (192.46)* | 224.08 (195.47)* |
| EPI 10 uM | 647.4 (232.08) | 405 (258.69)* | 449.47 (261.5)* |
| Collagen 2 mg/ml | 331.05 (65.56) | 238.41 (91.84)* | 260.76 (75.17)* |
| Collagen 5 mg/ml | 392.89 (51.82) | 344.48 (81.84)* | 362.57 (67.17)* |
| ADP 1 uM | 86.12 (89.21) | 0.54 (2.42)* | 2.15 (6.39)* |
| ADP 5 uM | 365.82 (74.24) | 11.82 (20.36)* | 19.87 (27.98)*<sup>‡</sup> |
| ADP 10 uM | 379.95 (40.63) | 36.18 (41.51)* | 57.59 (53.17)*<sup>‡</sup> |
| PFS | 0.59 (1.91) | -5.43 (1.83)* | -4.82 (1.75)* |
| Platelet count (10<sup>9</sup>/L) | 238.31 (71.29) | 244.14 (73.89)* | 254.41 (72.85)*<sup>‡</sup> |
| Mean platelet volume (fl) | 8.34 (0.73) | 8.29 (0.74)* | 8.35 (0.79) |
| Abbreviations per Supplemental Table 3. |  |  |  |

**Supplemental Table 5.**
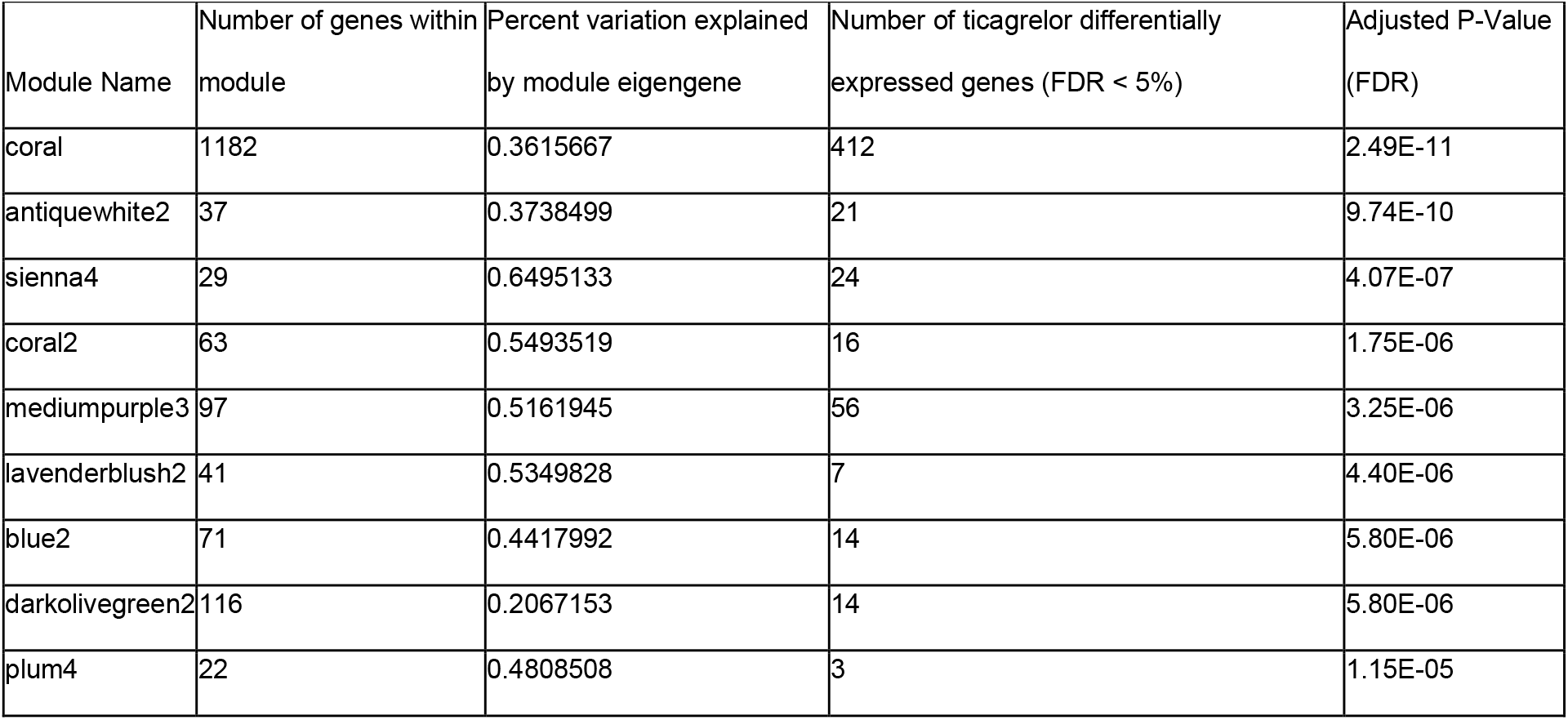
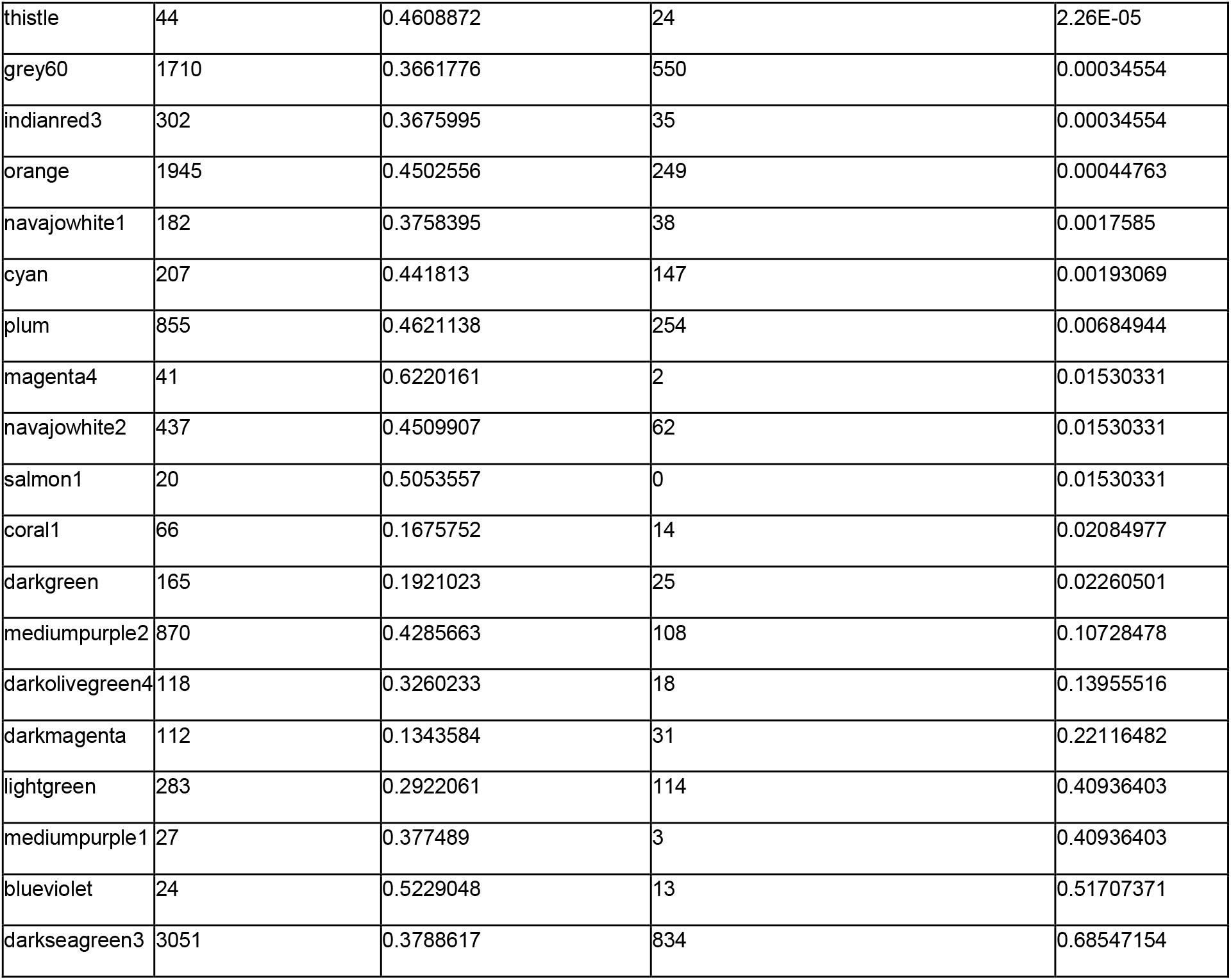
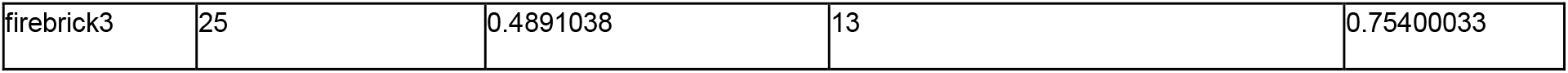
Attributes of WGCNA modules

## Acknowledgements

We would like to thank Ms. Pamela Isner and Dr. Thomas Burke in the Duke Center for Applied Genomics &Precision Medicine for their technical assistance with biobanking and sample processing.

