## Supplementary figures and images for "Ticagrelor responsive platelet genes are associated with platelet function and bleeding"

### Supplemental Figure 1

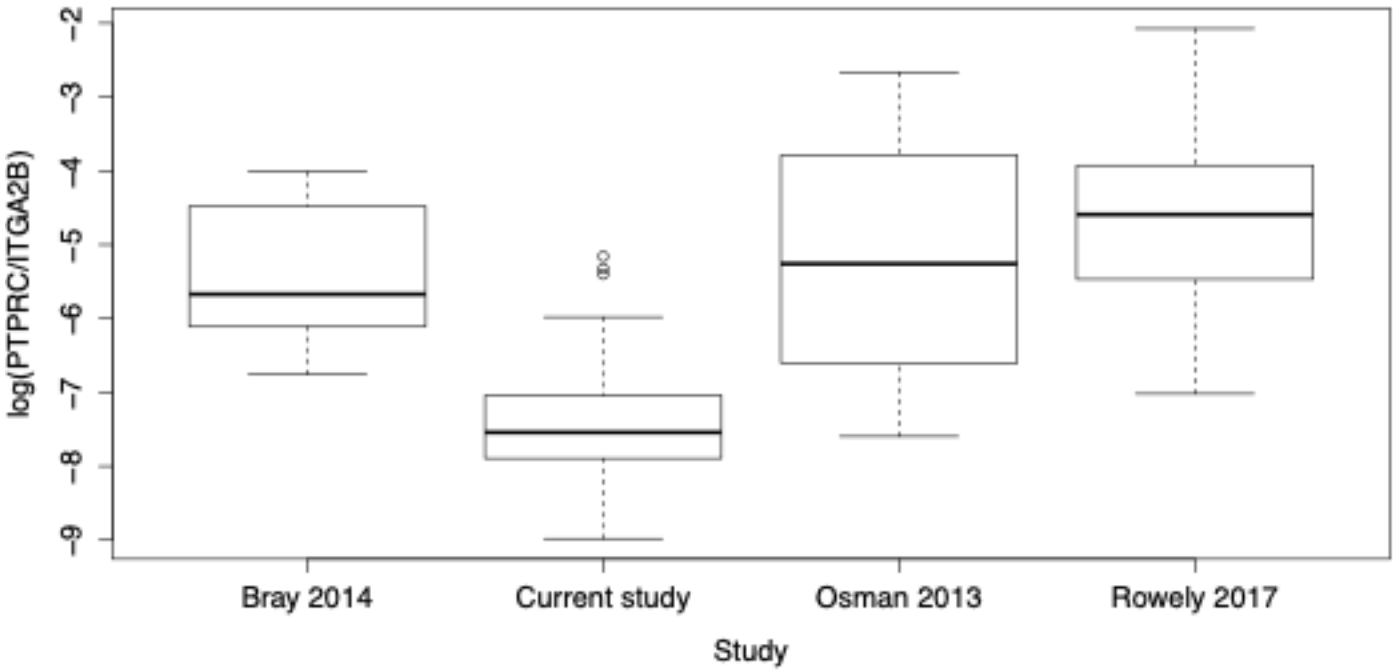

### Supplemental Figure 2

**Clustering of module eigengenes**

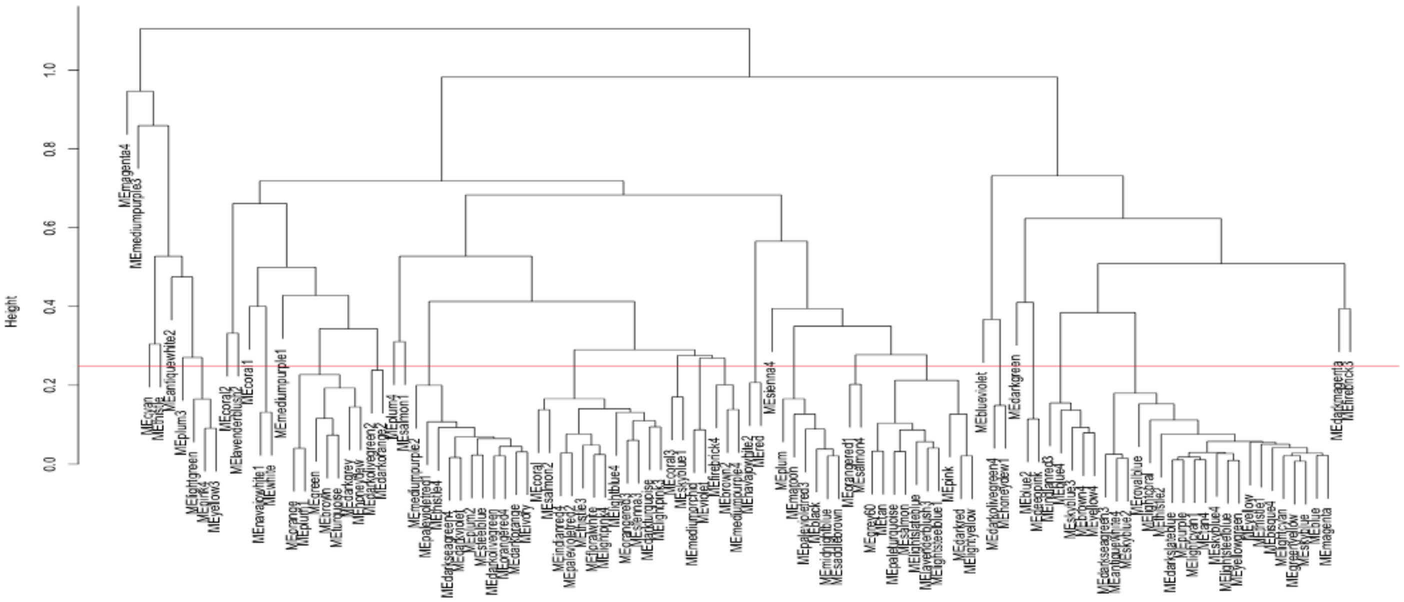
